# SARS-CoV-2 genome diversity at the binding sites of oligonucleotides used for COVID-19 diagnosis

**DOI:** 10.1101/2020.12.10.20236943

**Authors:** Renan Valieris, Michał B. Kowalski, Alina Frolova, Witold Wydmański, Johnathan Foox, Giovana T. Torrezan, Ewelina Pośpiech, Wojciech Branicki, Kasthuri Venkateswaran, Bharath Prithiviraj, Ramasamy Dhamodharan, Klas I. Udekwu, Diana N. Nunes, Dirce M. Carraro, Christopher Mason, Paweł P. Łabaj, Israel Tojal da Silva, Emmanuel Dias-Neto

## Abstract

**Importance:** SARS-CoV-2 genomic variants impacts the overall sensitivity of COVID-19 diagnosis, leading to false-negative diagnosis and the continued spread of the virus.

**Objective:** To evaluate how nucleotide variability in target primer binding sites of the SARS-CoV-2 genomes may impact diagnosis using different recommended primer/probe sets, as well as to suggest the best primer/probes for diagnosis.

**Design:** We downloaded 105,118 public SARS-CoV-2 genomes from GISAID (Sept, 25th, 2020), removed genomes of apparent worst quality (genome length <29kb and/or >5% ambiguous bases) and missing metadata, and performed an analysis of complementarity for the 13 most used diagnostic primers/probe sets for RT-PCR detection. We calculated the N rate and % of genome recovery, with all primer/probe-sets considering viral origin and clade. Results: Our findings indicate that currently, the Paris_nCoV-IP2, -IP4 and WHO|E_Sarbeco primer/probe sets for COVID-19, to perform the best diagnostically worldwide, recovering >99.5% of the good quality SARS-CoV-2 genomes from GISAID, with no mismatches. The Chinese_CDC|2019-nCoV-NP primer/probe set, among the first to be designed during the pandemic, was the most susceptible to currently most abundant SARS-CoV-2 variants. Mismatches encompassing the binding sites for this set are more frequent in Clade-GR and are highly prevalent in over 30 countries globally, including Brazil and India, two of the hardest hit countries. **Conclusions:** Detection of SARS-CoV-2 in patients may be hampered by significant variability in parts of the viral genome that are targeted by some widely used primer sets. The geographic distribution of different viral clades indicates that continuous assessment of primer sets via sequencing-based surveillance and viral evolutionary analysis is critical to accurate diagnostics. This study highlights sequence variance in target regions that may reduce the efficiency of primer:target hybridization that in turn may lead to the undetected spread of the virus. As such, due to this variance, the Chinese_CDC|2019-nCoV-NP-set should be used with caution, or avoided, especially in countries with high prevalence of the GR clade.

**Key Points:** 

**Question:** How variable are the binding-sites of primers/probes used for COVID-19 diagnosis?

**Findings:** We investigated nucleotide variations in primer-binding sites used for COVID-19 diagnosis, in 93,143 SARS-CoV-2 genomes, and found primer sets targeting regions of increasingly nucleotide variance over time, such as the Chinese_CDC|2019-nCoV-NP. The frequency of these variations is higher in Clade-GR whose frequency is increasing worldwide. Paris_nCoV-IP2, IP4 and WHO|E_Sarbeco performed best.

**Meaning:** We suggest the use of some sets to be halted and reinforce the importance of a continuous surveillance of SARS-CoV-2 variations to prompt the use of the best primers.

## Introduction

The current COVID-19 pandemic that resulted from the global spreading of SARS-CoV-2 has shown the importance of fast access to reliable viral detection methods. Indeed, viral containment measures can only be effective through the fast, broad, and accurate identification of subjects who carry active infection and may be actively spreading SARS-CoV-2. In this sense, the identification and isolation of SARS-CoV-2 cases is one of the most effective means by which to halt the viral spread and to reduce the number of new cases of COVID-19 (1,2).

RT-qPCR using primer (and probe) sets that target selected regions of the viral genome is the current gold standard and most common diagnostic tool for SARS-CoV-2. Hence, for high specificity and to avoid false-negatives, genomic variants need to be taken into consideration for the primer/probe designs, such that they can avoid target variations that would restrict or inhibit primer-template interaction during amplification. Due to continuous evolutionary selection by targeting sites of the viral genome, as well as viral genome drift, the primers and probes used for SARS-CoV-2 detection, as well as for other related methods like Loop-mediated isothermal amplification (LAMP) assays or other similar approaches -- should be constantly revisited in light of emerging genetic variation and fixation.

Here we evaluated the binding sites of the primers/probes most commonly used for COVID-19 diagnosis, among SARS-CoV-2 genomes downloaded from the GISAID database (www.gisaid.org - as of Sept., 25th, 2020 - deposited since Dec. 2019). This enabled the investigation of viral genome evolution, diversity and variant-spreading during the COVID-19 pandemics. Our data suggests that the continued emergence of genomic variants in SARS_CoV-2 may increase false-negative rates and lead us to recommend that: i) the use of Chinese_CDC|2019-nCoV-NP-set should be immediately discontinued; ii) more than one primer/probe-set should be used to reduce the frequency of false-negatives; iii) SARS-CoV-2 genomic variability should be continuously assessed by sequencing as a means of constant monitoring variations that may affect diagnosis.

## Results

We investigated the capability of 13 of the most used primer/probe sets for COVID-19 diagnosis (**Table S1**), to recover SARS-CoV-2 genomes from GISAID considering first no mismatches and also a maximum of two mismatches. After the exclusion of poor quality genomes (<29Kb and/or >5% of ambiguous ‘N’ bases) or missing metadata, 93,143 genomes remained (88.6%) (**Fig. S1**). The places of origin and clades of these viral genomes were also considered and, in some cases, genomes from distinct locations were clustered (e.g. England and Scotland, merged as the United Kingdom). Also, only countries/clusters with at least 10 viral genomes available were evaluated.

The Chinese_CDC|2019-nCoV-NP set displayed acceptable recovery rates in a few countries from Asia (**Fig. 1a and Table S2**). Despite this, this set performs poorly in recovering most SARS-CoV-2 genomes from GISAID. When no mismatches are allowed, recovery rates below 60% were obtained for >30 countries on three different continents (South America, Europe and Asia), including countries with significant burden such as India and Brazil (**Fig. 1a**). Assuming that that two mismatches would still allow sensitive amplification, the picture is still worrisome for most regions of the world (**Fig**.**1b**), a finding that reflects expectant evolutionary changes in this region of the SARS-CoV-2 genome. We observe that the variants in the binding site of the Chinese_CDC|2019-nCoV-NP set are more frequent in the clade GR, allowing recovery rates (no mismatches) from 80.8% in North America to 33.9% in Oceania (**Table S2**). Clade O appears to concentrate variants in binding sites of US_CDC|2019-nCoV_N1 and US_CDC|2019-nCoV_N3 sets **(Fig. 1a, 1c)**.

**Figure 1:**
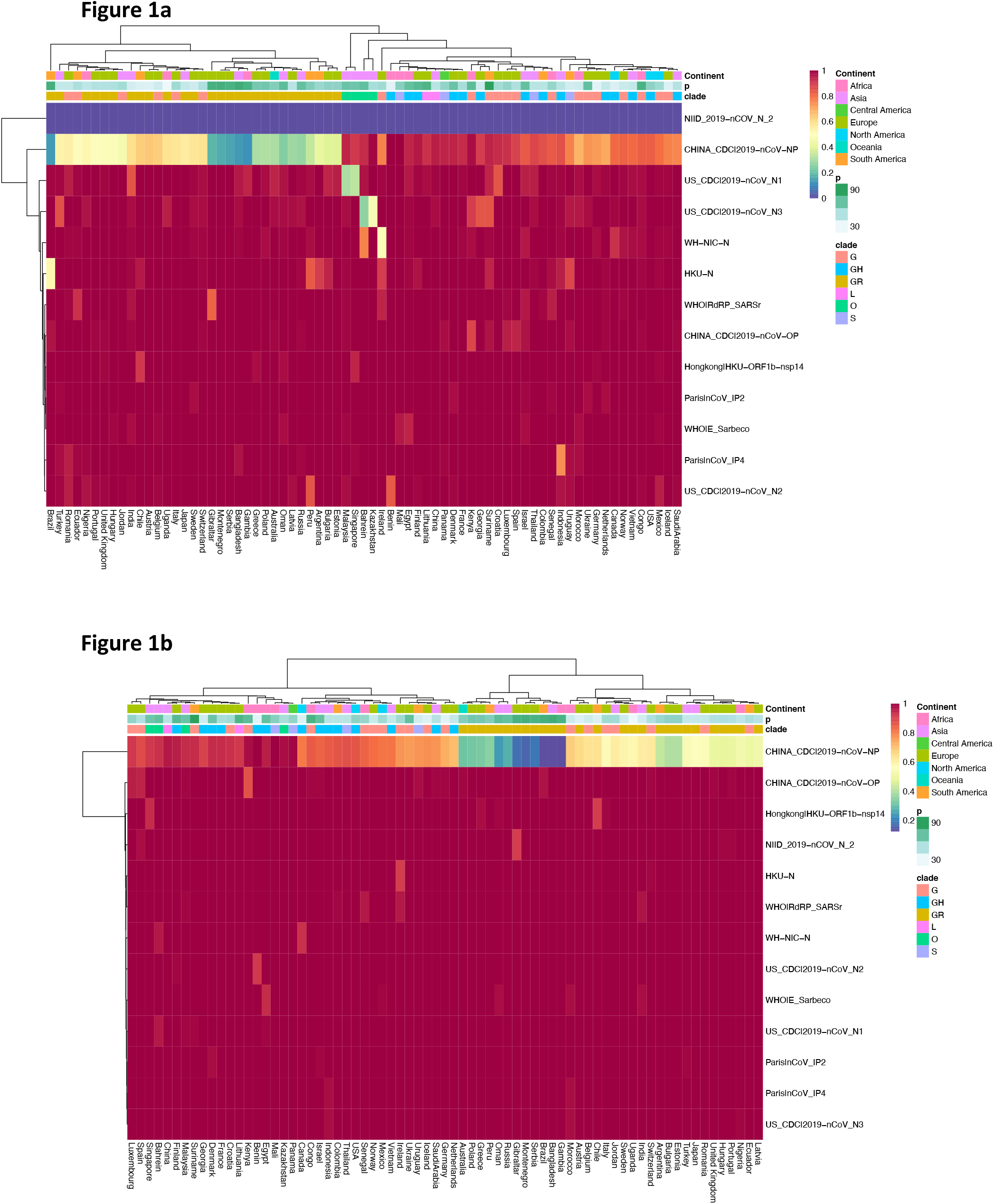
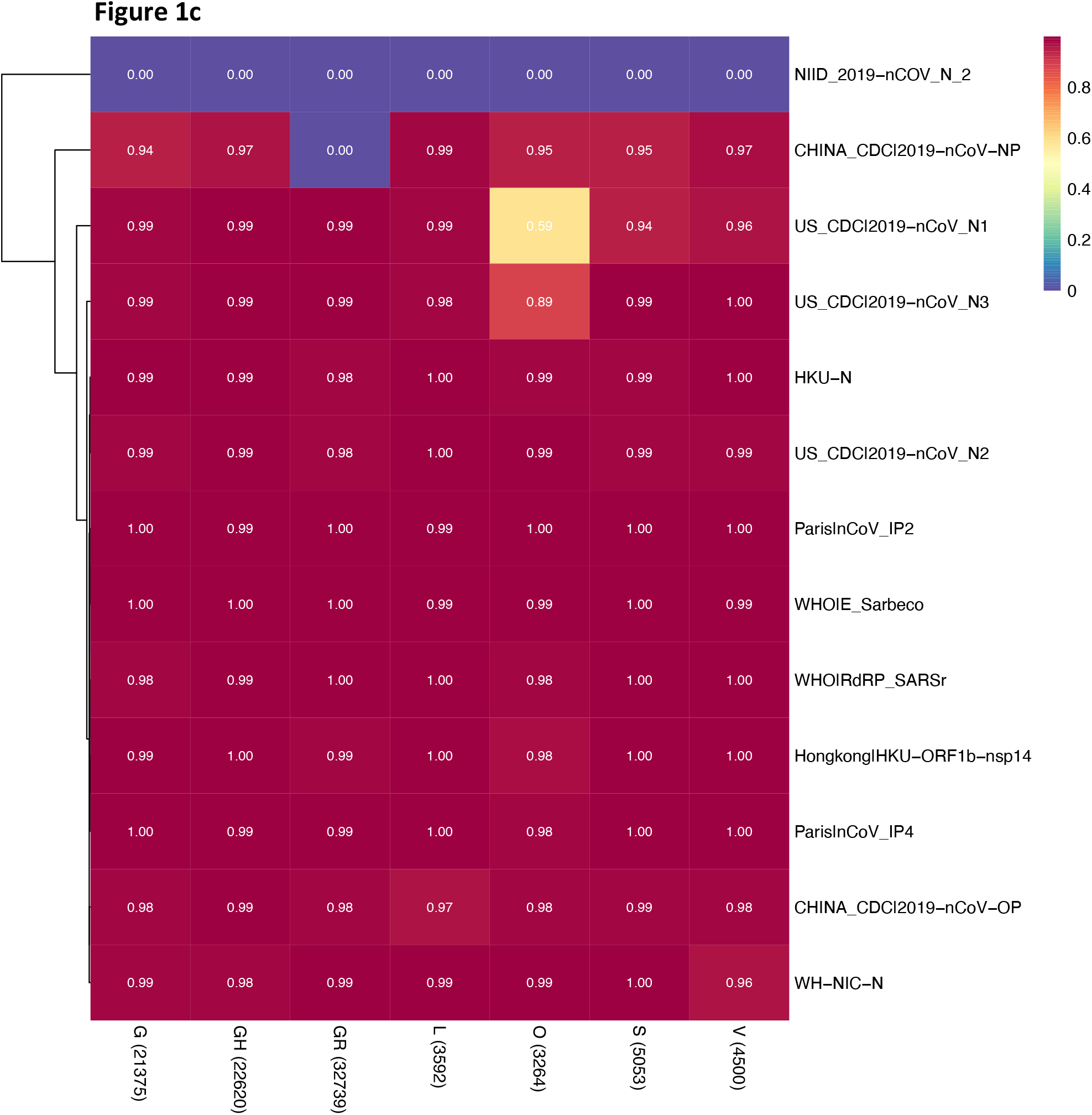
Heatmaps showing recovery rates of genomes from distinct countries of the world according to the presence of nucleotide variants in the binding sites of primers/probes and SARS-CoV-2 genomes. The recovery rate results when the requirement of a perfect complementarity between primers/probes for successful amplification is applied is shown in Fig. 1a, whereas recovery rates when up to two mismatches are allowed (Fig. 1b). Top colored lines indicate the continental location of the countries; the next two lines show the proportion of the predominant clade in a particular region (p) and the respective predominant clade (clades). Fig. 1c shows the efficacy of viral recovery (no mismatches allowed) for each viral clade and for each primer/probe set. Numbers inside parentheses indicate the number of genomes for a given clade. For the NIID_2019-nCOV_N_2 and WHO_RdRP_R sets, see Suppl. Information for typos and errors in published primer-sequences.

Other primer/probe sets also showed low recovery rates for specific regions, such as HKU-N for Brazil (total 6 variant positions distributed in primers F, R and the probe); US_CDC|2019-nCoV_N1 for Malaysia and Singapore (3 variant positions located in primer F and the probe); and US_CDC|2019-nCoV_N3 for Bahrain and Kazakhstan (7 variants for primers F, R and the probe) (**Table S3**). Caution should be used as the numbers of viral genomes in GISAID are tremendously biased towards Europe and the US, and the minimum of 10 genomes used here may not reflect actual viral diversity present in some countries.

Next, we investigated the temporal prevalence of each clade since the start of the pandemic. We charted significant fluctuations in clade prevalence and found SARS-CoV-2 clades G, GH and GR to have increased significantly relative to others, suggestive of positive selection of these variants **(Figure 2)**. These results however, can also be explained by reduced diagnostic accuracy as a consequence of the mismatched evolving target sites. Accordingly, genomic variations should be considered when primers/probes sets are to be selected.

**Figure 2.**
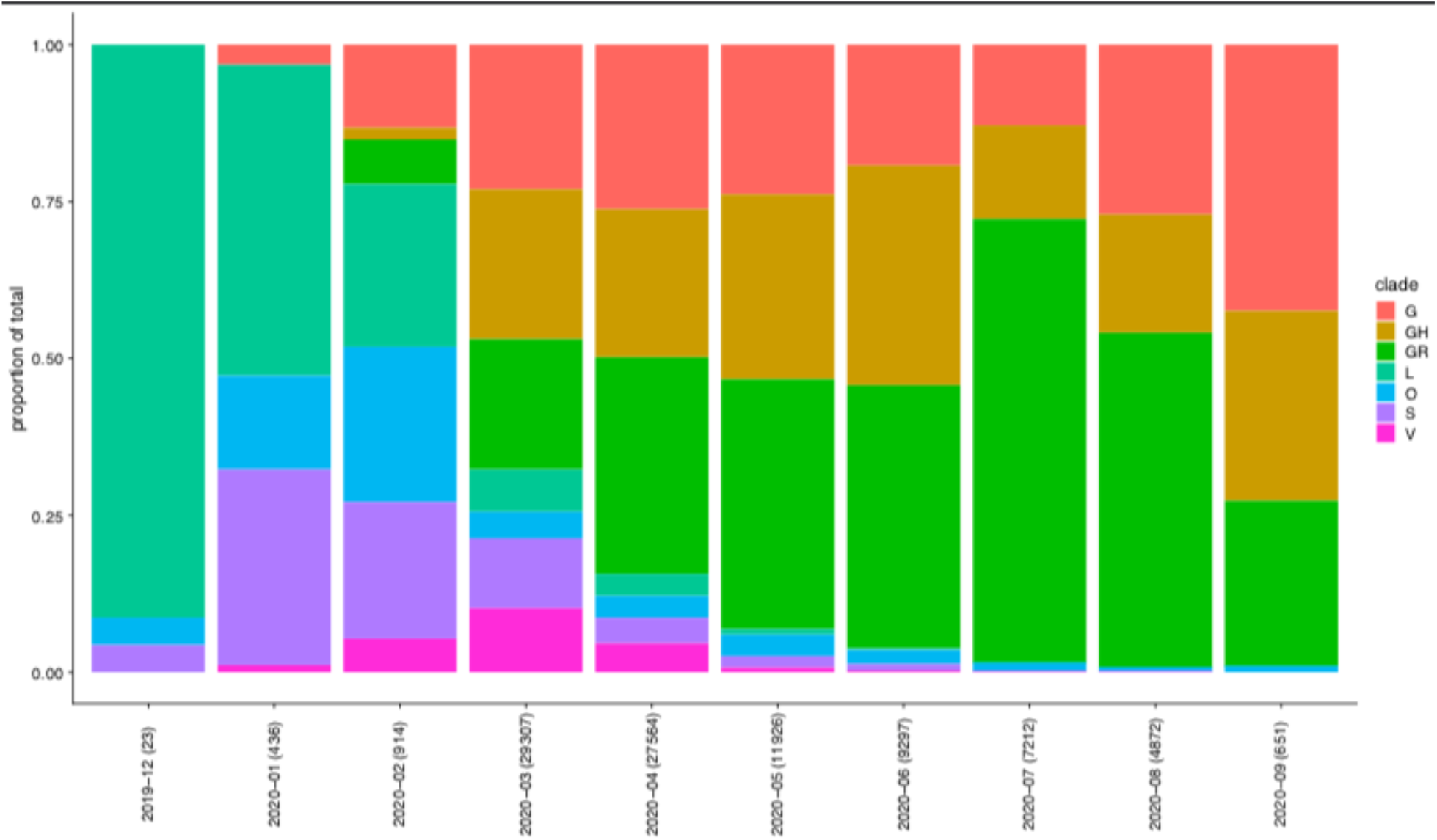
Percentage of SARS-CoV-2 clades in the world according to month when the sequences were deposited in GISAID. The number of genomes from each month are indicated (parenthesis). Genomes with no information about the month in which they were sequenced (∼1%) have not been considered.

When considering the top-three countries in the world in terms of number of COVID-19 cases (USA, India and Brazil - **Fig. S2**) we see that - despite the reduced number of genomes from Brazil - the proportion of viral clades varies significantly for all these 3 countries. This result highlights the importance of considering regional and temporal clade structures for selecting primer/probe sets for diagnosis.

As an exercise to evaluate variations in the binding sites from patient-derived samples that utilized RNA-sequencing to profile the virus, we also analyzed the shotgun sequencing of 926 COVID-19 positive samples from New York-Presbyterian and Weill Cornell Medicine patients, from New York City. Again, most mismatches were seen for the Chinese_CDC|2019-nCoV-NP set, but we have also observed variations in the binding sites for the reverse primers of the US_CDC|2019-nCoV_N2 and N3 sets as well as WH_NIC sets **(Fig. S3)**. The most frequent SARS-CoV-2 genomic alteration affecting the Chinese_CDC|2019-nCoV-NP corresponds to the 5’-end of the binding site of the forward primer (3) - a continuous stretch of three nucleotide substitutions (GGG→AAC). Viral genomes with this variation corresponded to ∼13% of the available SARS-CoV-2 genomes as of 22 March 2020 (3) but have now reached ∼64% of GISAID sequences. This variation found in 88% of a SARS-CoV-2 genome cohort comprised of 640 sequences from Indian patients and in all of 40 recently confirmed cases in São Paulo, Brazil. These findings provide multiple lines of evidence of these variations and their significant frequency in patient-derived samples. Whereas a scenario of high/medium viral load may not lead to false-negatives after diagnosis with this set (as the most frequent variant would affect the 5’ end of the F-primer), one possibility is that in cases of low viral loads or less efficient swabbing, Ct values may be shifted above the detection threshold, leading to false-negatives and a consequent spreading of this variant. While Vogels et al. 2020 (3), argue that the precise location of this mismatched sequence may not impact COVID-19 diagnosis, the hypothesis remains to be tested.

The continued evolution of the virus may result in more variants in these binding sites and other genome regions and, in this case, the continued use of the Chinese_CDC|2019-nCoV-NP primer-set may increase false-negatives rates. As can be seen from **Table S3** other variants in the binding sites of F and R primers of the Chinese_CDC|2019-nCoV-NP set have also been detected in our analysis. Furthermore, it is relevant to document that 2/40 cases sequenced in Brazil showed two additional substitutions, one detected at the 7^th^ nt from the 5’end (C>A) and the other at the 4^th^ nt from the 3’end (G>T). These changes, which may have stronger impacts on primer efficacy – especially if combined with the more upstream mismatches, indicate that nucleotide substitutions continue to accumulate in this genome region. Importantly, none of these two extra variations have been described in this GISAID-version, further reinforcing the need to a continuous effort on cataloguing viral genome sequences.

As the identification of subjects carrying SARS-CoV-2 is in itself an important barrier to the propagation of the virus, we speculate that the increased global spread of viruses carrying this variant may be a consequence of using primer/probe sets that fail to properly identify positive cases. It is worth mentioning the early use of the Chinese_CDC|2019-nCoV-NP primer set in earlier manuscripts (4-7). While mass screening is needed to control the spread of the disease, the lack of proper detection of viruses carrying some variants will not only result in problems at individual/community level as well as on a global scale. Moreover, such missed diagnoses could contribute to the ongoing spread of the virus, increasing the number of new cases and deaths from COVID-19, and lead to continued pandemic spread due to misdiagnoses.

The variations seen here may impact COVID-19 diagnosis by RT-PCR, but may also impact other diagnostic approaches such as LAMP (8) and the Ion AmpliSeq™ SARS-CoV-2 Research Panel. There we observe that a synonymous variation (c.14143C>T; p.Leu4715Leu; orf1ab) in the binding site of the ORF1AB primer resulted in decreased coverage of this amplicon by 1-2 orders of magnitude (16/25 samples from a Polish clinical cohort). An analysis of GISAID genomes showed this variant to be present in 10% of Polish sequences and in about 17% of sequences from other European countries as well as other continents (**Fig. S4**)

Conversely, our study also revealed primers that can currently be used with confidence. Using current data, we found that Paris_nCoV-IP2 and -IP4, and WHO|E_Sarbeco have shown the best performance in terms of full match to SARS-CoV-2 genomes worldwide, all capturing above 99.5% of the good quality SARS-CoV-2 viruses (**Table S4**). Importantly, geographic variations need to be considered and monitored overtime, despite the good performance of these primers at this time.

Although the currently observed genome variations would not always impact SARS-CoV-2 detection, since partial amplification can still occur, we propose: i) the use of more than one primer/probe set to minimize false-negative rates; ii) the use of the Chinese_CDC|2019-nCoV-NP set to be discontinued; iii) the permanent sequencing surveillance of SARS-CoV-2 genome around the world, especially from non-primer-biased, environmental samples - allowing viral genome variant monitoring and the careful selection of the best primers/probes as a means to reduce false-negatives and disease spreading.

## Data Availability

The raw data set used in this paper can be download at https://www.gisaid.org/

## Acknowledgements

DMC and ED-N acknowledge Conselho Nacional de Pesquisas (CNPq – Brazil). ED-N is thankful for the support received from Associação Beneficente Alzira Denise Hertzog Silva (ABADHS, Brazil).

## Supplementary Information

### Supplementary Note

The WHO_RdRP_R primer contains an error in a degenerate base at position 12 (instead of S - C/G – it is certainly a T; see (S1), leading to zero-recovery in our analysis (Fig. 1a). The correction of this mistake would allow for recovery of 91.19% of GISAID genomes (Fig. S-MCB.1 & S-MCB.2; see online material for detailed analysis). Extra attention should be given to this as the wrong primer sequence is still widely found in the literature.

The NIID_2019-nCOV_N_R2 primer appears to have an error/typo even in the erratum where it is indicated that “The reverse primer (NIID_2019-nCOV_N_R2) sequence should be replaced with TGGCAGCTGTGTA**<u>G</u>**GTCAAC. The corrected nucleotide is bold and underlined.” However, our analysis suggests there to be another typo and the correct primer sequence should be TGGCA**<u>C</u>**CTGTGTAGGTCAAC (with the previously wrong base marked in bold and underlined). In order to highlight this discrepancy we performed our searches according to the erratum (S2).

### Bioinformatics

Genomes, primers and probes evaluation analyses were performed by running in-house pipelines. We first downloaded all SARS-CoV-2 RNA sequences (N=105,118) available at GISAID as of September 25th. After removal of Ns from up and downstream of each virus genome sequence, only genome sequences with at least 29Kb and a maximum of 5% of N (ambiguous bases) were used for the alignment of primers and probes. These sequences (N=13 primer/probe sets) were matched against the filtered genomic sequences with the software bowtie2 (v2.3.5.1) (S3); command line options: --end-to-end --very-sensitive -a) in paired-end alignment mode. After, probes sequence from each pair of primers were mapped to each virus sequence by bowtie2 and then merged with the primer alignments results. Exact proper pair alignments allowing up to 2 mismatches outside of the last 5bps of the 3’ end of each primer were considered for further analyses.

**Table S1.**
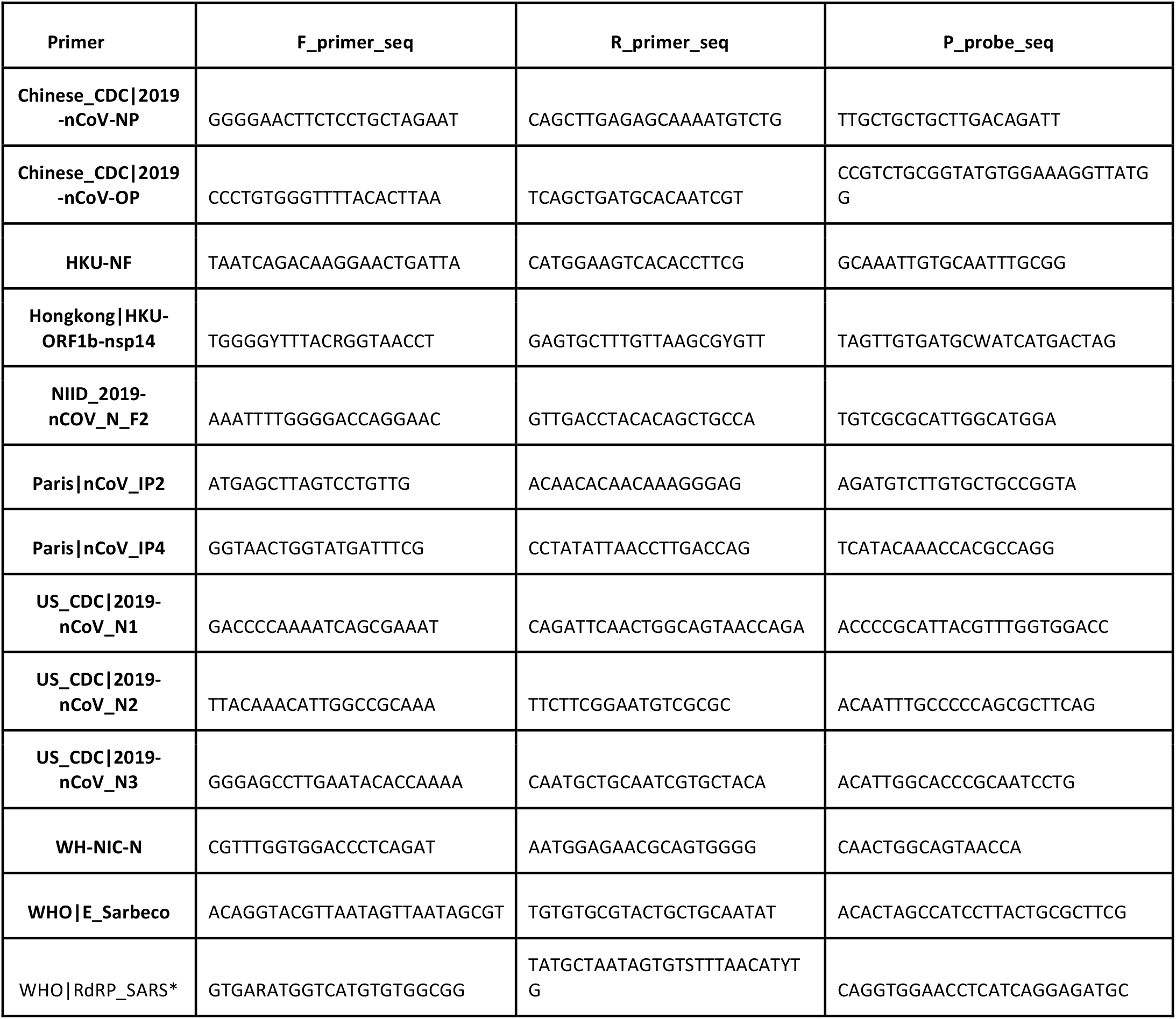
Primers and probes evaluated.

**Table S2.**
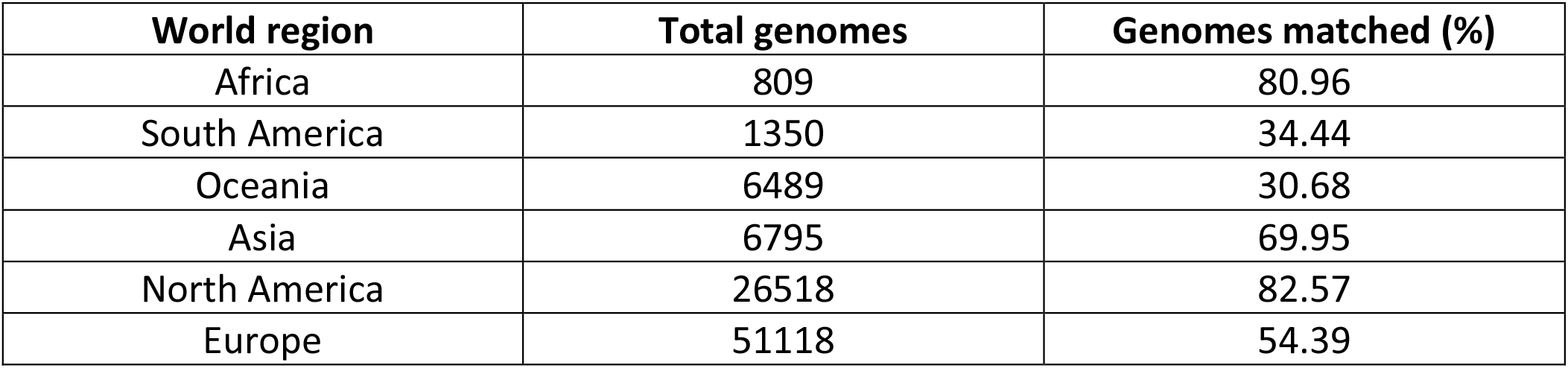
Chinese_CDC|2019-nCoV-NP primers/probe set performance in the different world regions with zero mismatches allowed. The percentage of matching sequences were calculated based on the number of amplicons of non-zero length.

| World region | Total genomes | Genomes matched (%) |
| --- | --- | --- |
| Africa | 809 | 80.96 |
| South America | 1350 | 34.44 |
| Oceania | 6489 | 30.68 |
| Asia | 6795 | 69.95 |
| North America | 26518 | 82.57 |
| Europe | 51118 | 54.39 |

**Table S3.** Variations found for SARS-CoV-2 genomes from the GISAID database at each nucleotide base (from 5’- to 3’-end) of the most varying primer/probe sets according to Figure 1a. Numbers in each cell represent how many genomes carry that specific nucleotide of the indicated primers or probes.

**Table S4.** Genome recovery rates for different primers and probes, with and without mismatches.

| Primer-set | Genomes recovered with no mismatches |  | Genomes recovered with up to two mismatches |  | Bad hits<sup>1</sup> |
| --- | --- | --- | --- | --- | --- |
|  | F+R | F+R+probe | F+R | F+R+probe |  |
| NIID_2019-nCoV_N | 0 | 0 | 92950 | 92918 | 199 |
| CHINA_CDC\|2019-nCoV-NP | 57859 | 57794 | 59366 | 59337 | 312 |
| US_CDC\|2019-nCoV_N1 | 92521 | 90560 | 92956 | 92935 | 219 |
| CHINA_CDC\|2019-nCoV-OP | 92090 | 91769 | 92332 | 92181 | 182 |
| US_CDC\|2019-nCoV_N2 | 92242 | 91893 | 93091 | 93055 | 67 |
| US_CDC\|2019-nCoV_N3 | 92296 | 91958 | 93111 | 93086 | 68 |
| WH-NIC-N | 92140 | 91958 | 92863 | 92848 | 303 |
| HKU-NF | 92258 | 92009 | 93064 | 93023 | 96 |
| Hongkong\|HKU-ORF1b-nsp14 | 92486 | 92268 | 92826 | 92608 | 246 |
| WHO\|RdRP_SARSr | 92639 | 92418 | 93006 | 92982 | 113 |
| Paris\|nCoV_IP4 | 92765 | 92477 | 93089 | 93053 | 103 |
| WHO\|E_Sarbeco | 92961 | 92748 | 93076 | 92985 | 114 |
| Paris\|nCoV_IP2 | 92865 | 92778 | 93012 | 92991 | 152 |

**Note:** The total number of genomes investigated is 93,143. The table has been ordered according to the total number of recovered genomes, with no mismatches for primers F+R and the probe. Bad hits<sup>1</sup> indicates genomes with mismatches corresponding to the very 3' end of the primers or probes, which shall preclude the proper amplification of the target.

## Suppl. Figures

**Figure S1.**
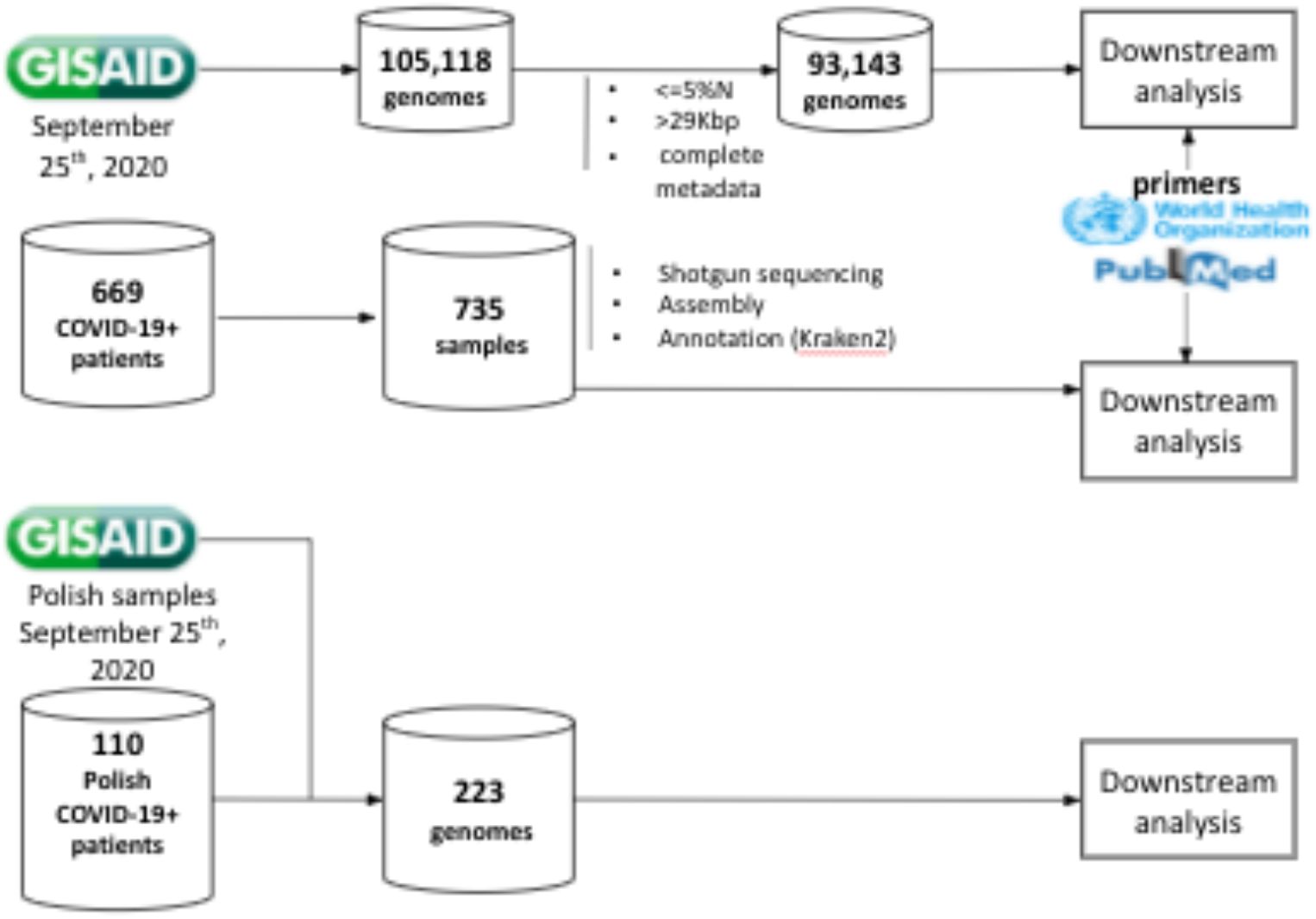
Analysis pipeline.

**Fig. S2a.**
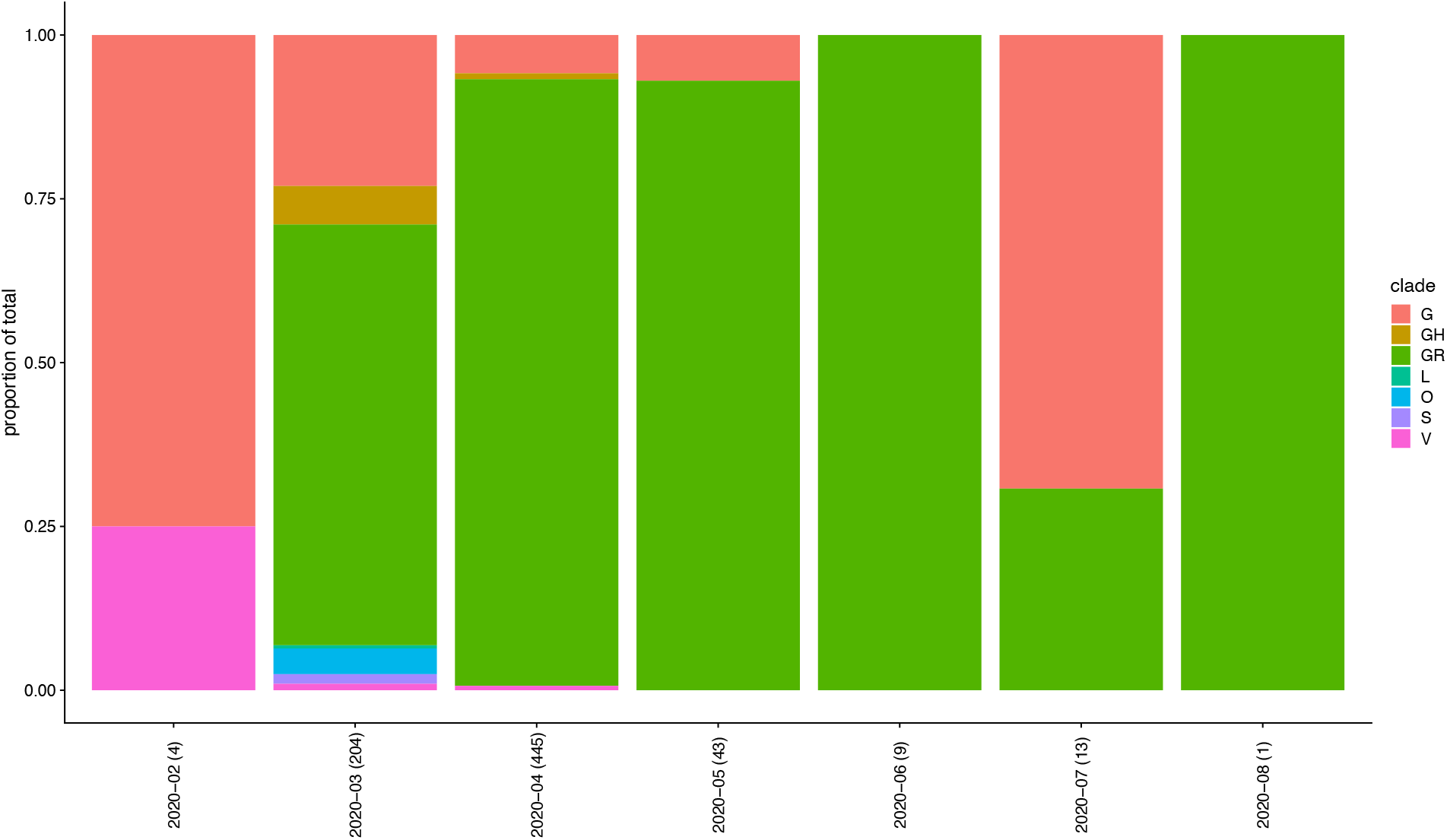
Brazil.

**Fig. S2b.**
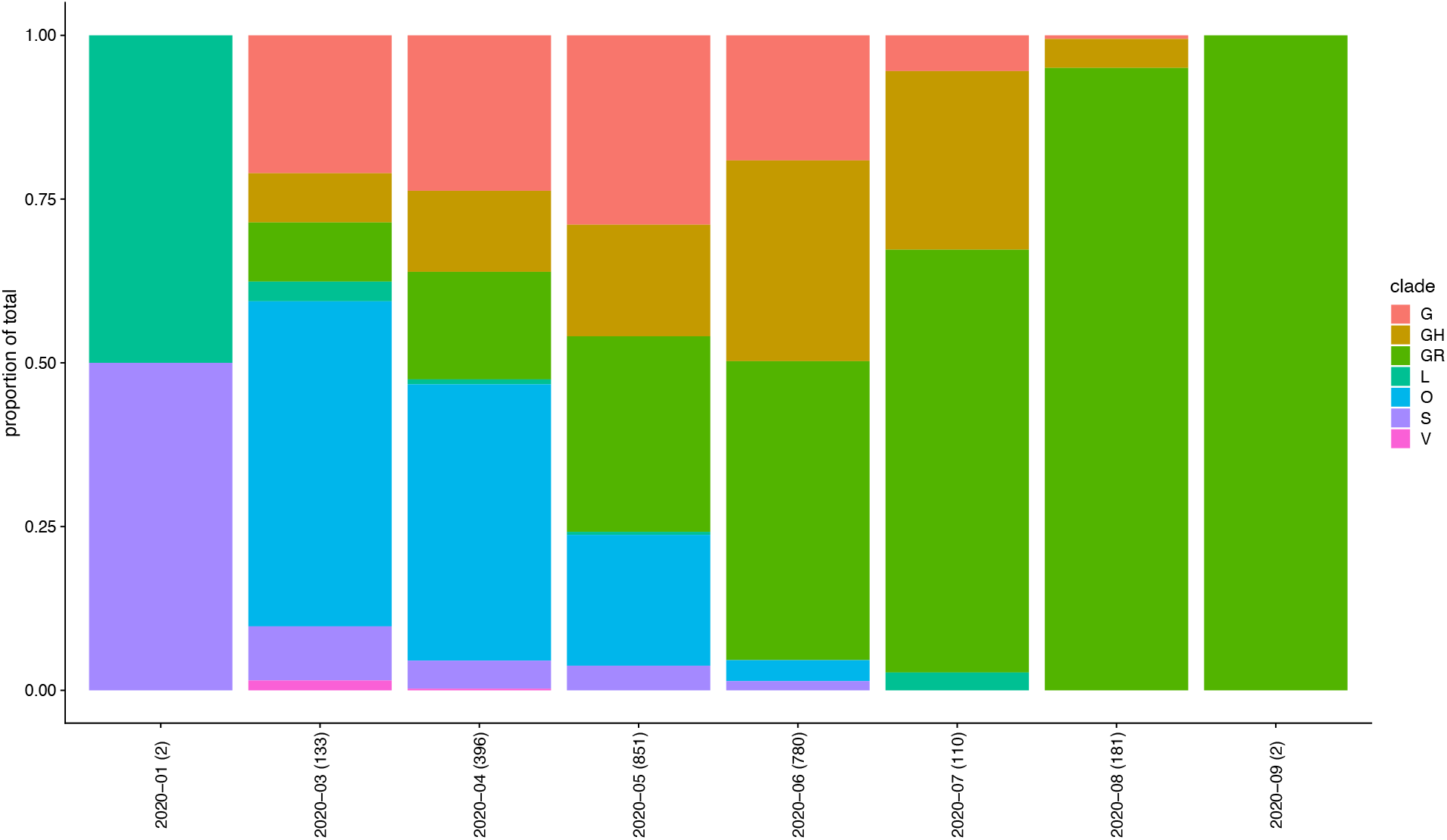
India.

**Fig. S2c.**
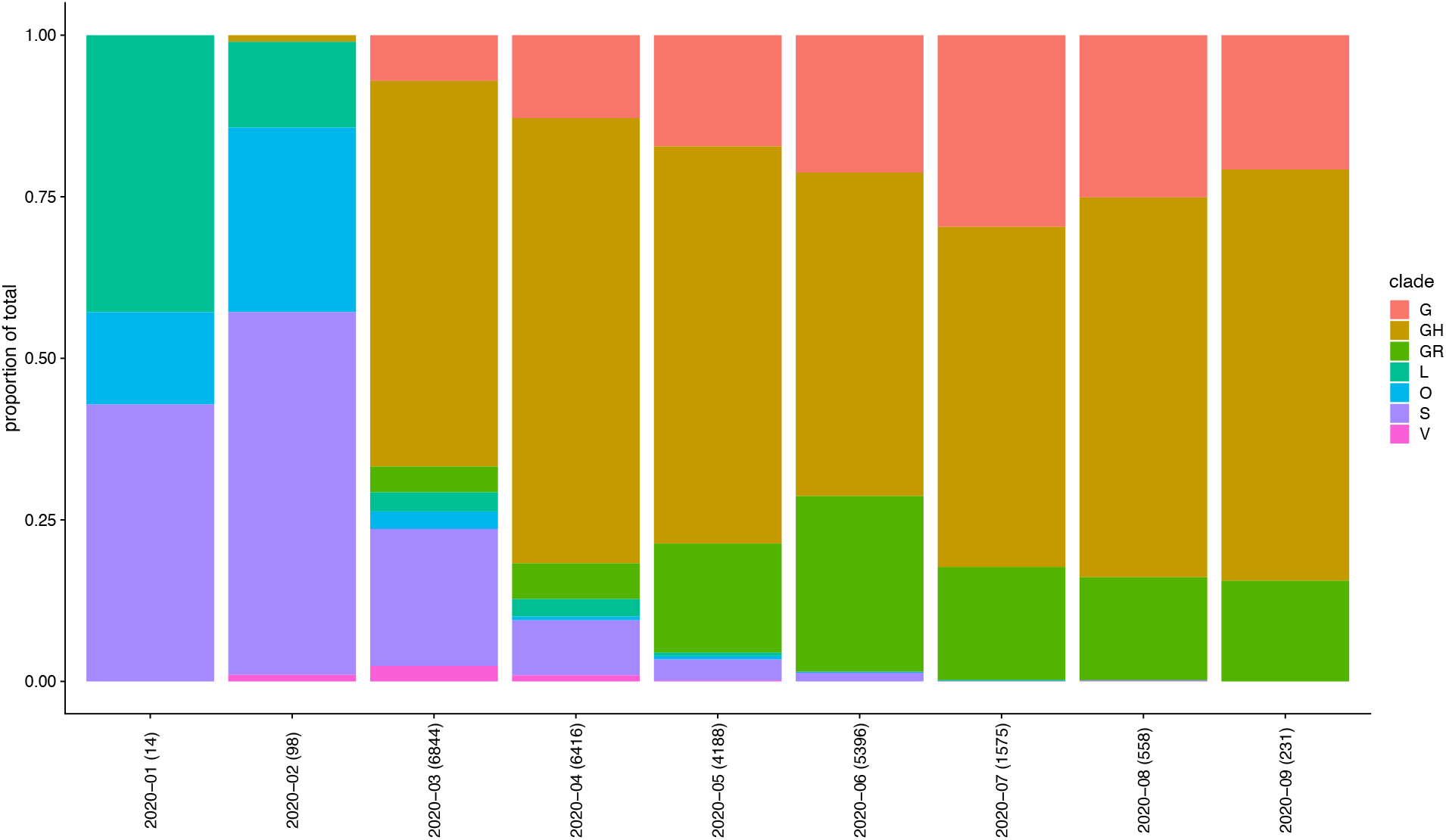
USA.

**Fig. S3.**
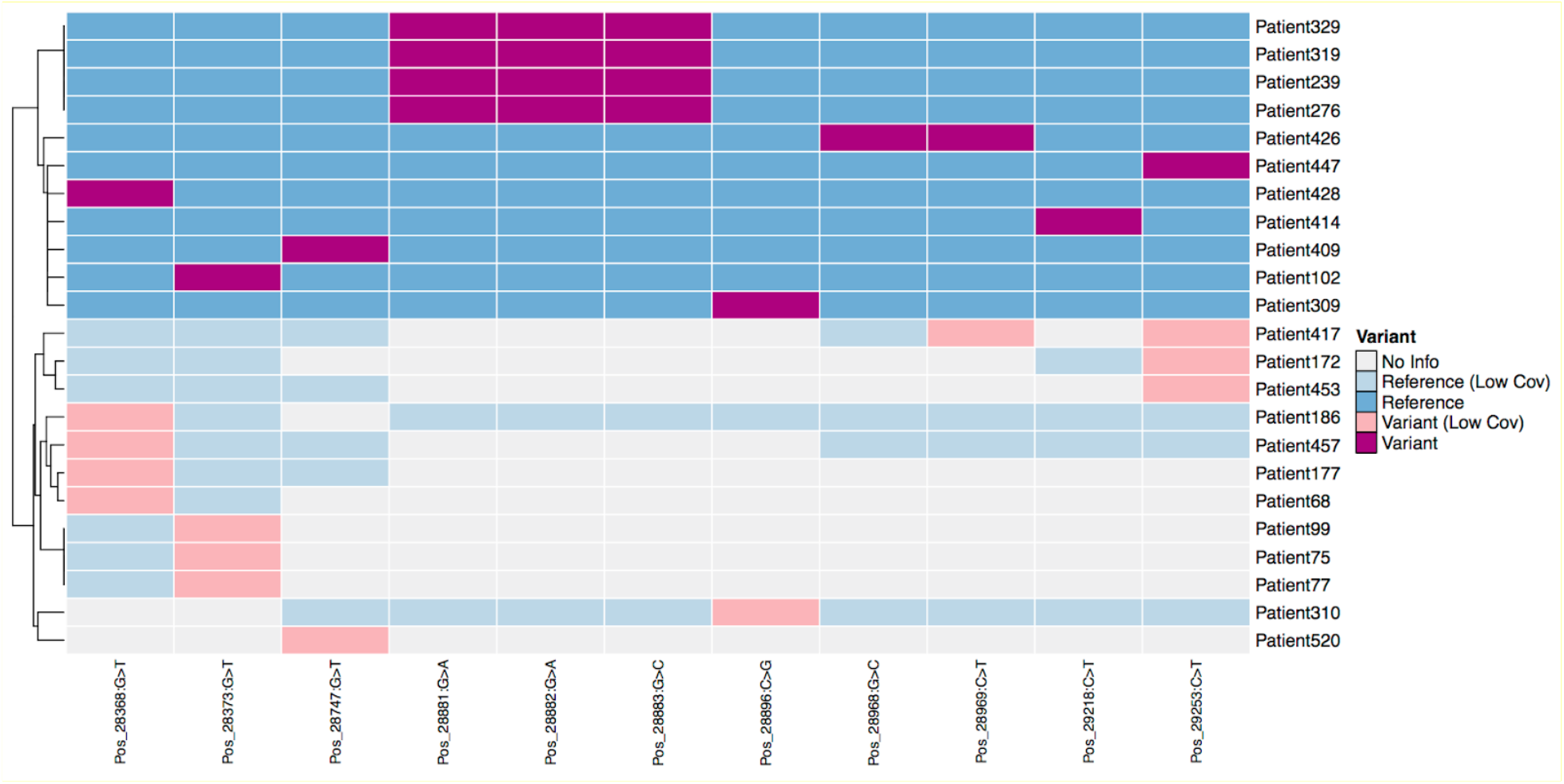
Presence of variations in primer-probe regions found from nasopharyngeal (NP) swab data from New York-Presbyterian and Weill Cornell Medicine patients. Total RNA was sequenced and viral reads were isolated and assembled from 926 NP swabs (see Butler *et al*., submitted 2020). Eleven variant alleles across binding sites for 13 primer/probe-sets were detected in viral genomes isolated from patient samples, including patients who exhibited variants in the coordinates corresponding to binding sites of the Chinese_CDC|2019-nCoV-NP set including the 3bp stretch of the forward primer (pos 28881-28883) and other variations along the binding site of this same forward primer (28896) as well as its reverse primer (28968 and 28969), along with variants in the binding sites for reverse primers of US_CDC|2019-nCoV_N2 (29218), US_CDC|2019-nCoV_N3 (pos 28747), WH-NIC-N (28373) and the vicinity of other primers/probes. Color shades correspond to depth of sequencing at each site (low coverage indicates <= 10 reads covering that site).

**Figure S4.**
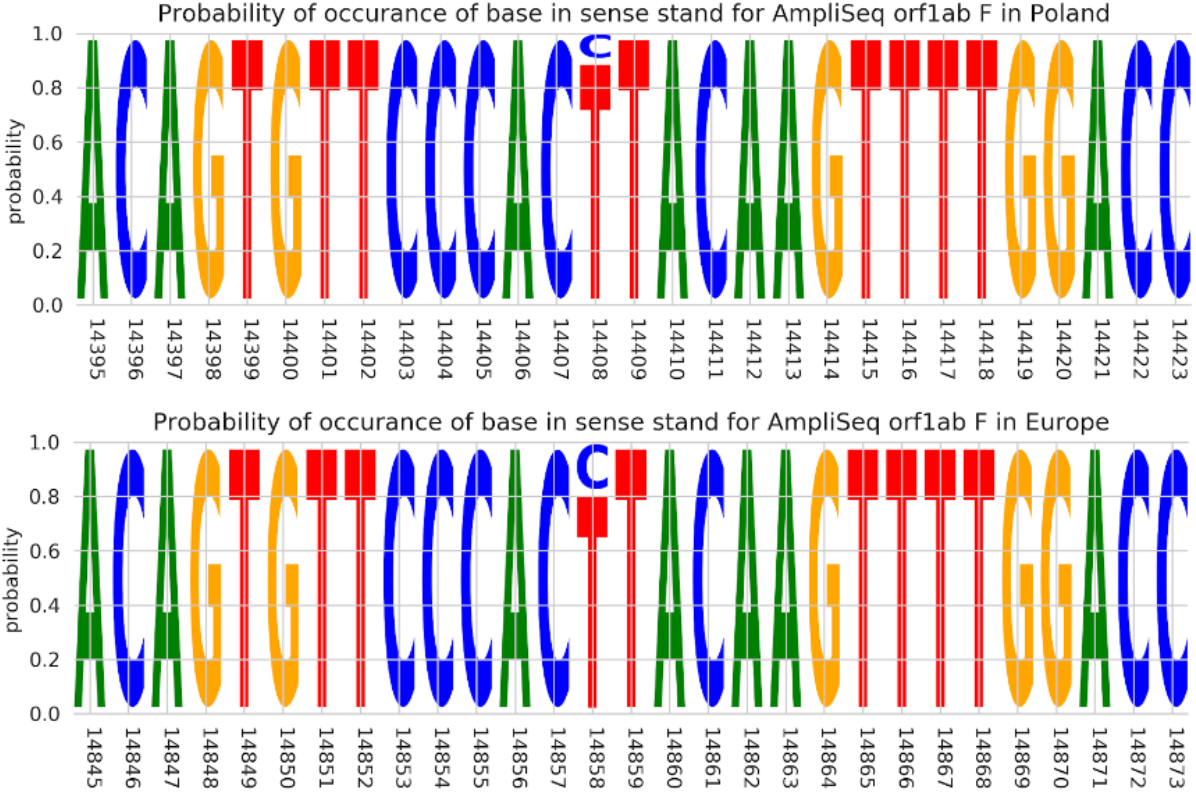
Variation in primer binding site of AmpliSeq™ SARS-CoV-2 Research Panel - c.14143C>T; p.Leu4715Leu; localized in ORF1ab. Top panel shows frequencies for Polish sequences with the 14408-variation of multi-aligned coordinates of 225 (110 from MCB, 115 from GISAID) Polish sequences. Bottom panel shows frequencies for other European countries with the variation in question localized on position 144858 of multi-aligned coordinates of 52983 European sequences. Variation frequencies found in other continents follow the same European pattern.

